# Health behaviours and their determinants among immigrants residing in Italy

**DOI:** 10.1101/2022.03.14.22272345

**Authors:** Giovanni Minchio, Raffaella Rusciani, Giuseppe Costa, Giuseppe Sciortino, Teresa Spadea

**Author notes:** **Corresponding author:** Giovanni Minchio, Department of Sociology and Social Research, University of Trento, Via Giuseppe Verdi, 26, 38122 Trento, Italy. **Authors’ contributions** GM: data curation, formal analysis, methodology, writing – original draft, writing – review & editing; RR: data curation, formal analysis, writing – original draft, writing – review & editing; GC: conceptualization, writing – review; GS: funding acquisition, project administration, writing – review; TS: conceptualization, methodology, supervision, writing – original draft, writing – review & editing. All the authors could access and verify the data, approved the manuscript and are responsible for the views expressed in it.

## Abstract

**Background:** The mechanisms that influence the uptake of risky behaviours among immigrants are influenced by the interrelation between characteristics operating in different phases of their migratory experience. Characterizing their behavioural risk profile is needed to prioritize actions for prevention and health services organization. We therefore analysed health behaviours and their determinants among immigrants in Italy, jointly accounting for sociodemographic factors, migration pathways and integration indicators.

**Methods:** Data come from a national survey conducted in 2011-2012 on a sample of about 12000 households with at least one foreigner residing in Italy. The independent impact of a variety of sociodemographic, migratory and integration characteristics on obesity, smoking and daily alcohol consumption was assessed using multivariable Poisson models.

**Results:** The survey involved more than 15,000 first generation immigrants. Unhealthy lifestyles are more common among men than among women and vary widely by ethnic group. There is a significant impact of employment status and family composition, while the educational level loses importance. Longer duration of residence and younger age at arrival are associated with an increased behavioural risk. Among women we also observed an independent impact of the integration indicators, less important for men.

**Conclusions:** The profile of the main unhealthy lifestyles among migrants is shaped by cultural, socioeconomic and migratory characteristics, which differ by gender. Understanding these factors can help to design tailored preventive messages, necessary to interrupt the deterioration of migrants’ health capital. Low levels of integration have an additional negative impact on health, so inclusion and integration policies should complement health promotion strategies.

## Introduction

A consistent body of literature suggests that the overall mortality risk of those who adopt healthy lifestyles could be lowered by 66% compared to people who engage in harmful lifestyles, such as smoking, being overweight or obese, having an unhealthy diet or not doing physical activity [1]. Characterizing the behavioural risk profile of a population is therefore essential in order to prioritize actions for prevention and health services organization. This is even more important among migrants. It is well known, in fact, that most migrants start moving from their countries when they are in good health conditions and therefore exhibit on average a lower mortality and morbidity than the host population (the so-called ‘healthy migrant effect’) [2]. However, recent studies also show that the profile of the main unhealthy behaviours among migrants is less favourable than what is generally observed for mortality or morbidity [3]. Therefore, the promotion of healthy behaviours among migrants would be necessary to interrupt the deterioration of their health capital and to prevent future poor health outcomes [4].

The mechanisms that influence the uptake of risky behaviours among immigrants are very complex, operating in different phases of their life course, going from reflecting past exposures to the present social disadvantage in working and living conditions, which they share with the lowest socioeconomic classes of the host population [5, 6]. Mechanisms are influenced by the interrelation between three broad groups of health determinants. Firstly, individual and contextual cultural and socioeconomic factors, which depends on both countries of origin and destination. Secondly, migration characteristics, such as age at arrival and length of residence, which affect the acculturation process [7]. Finally, the level of inclusion and integration in the new country as opposed to experiences of racial discrimination and legal constraints [8, 4]. The scientific literature on health-related lifestyles among immigrants, however, does not systematically account for these complex mechanisms. Furthermore, to our knowledge, no study analysing these mechanisms has been conducted specifically in Italy.

In the last decade, the share of immigrants in Italy has stabilized at around 8-9% of the total resident population, exceeding five million inhabitants. In 2011-2012, the National Institute of Statistic (Istat) conducted the first national survey on ‘Social conditions and integration of foreign citizen in Italy’ (https://www.istat.it/en/archivio/191097), investigating all aspects of individuals’ migration history, including an extensive set of questions on family, working and social life. The survey provided a unique opportunity for an in-depth analysis of health behaviours and their determinants, jointly accounting for sociodemographic factors and characteristics of the migration pathway and the integration process.

## Materials and methods

### The study population

We used data from the Istat multipurpose survey ‘Social conditions and integration of foreign citizen in Italy’, conducted in 2011-2012 on a representative sample of about 12000 households with at least one foreigner resident in Italy. We selected all the individuals born abroad and with foreign citizenship at birth (first generation of immigrants). Health behaviours were enquired only among people aged 15 years or more and we further decided to set an upper age limit to 64 years, both because the proportion of older foreigners was very small (about 3%) and to reduce the possible impact of the salmon bias, suggesting a selective return to countries of origin of older and sicker migrants [9]. The final study population consisted of 6947 men and 8783 women.

### Health behaviours

The health behaviours investigated in the survey were obesity, smoking and daily alcohol consumption.

Obesity is derived from the body mass index (BMI) calculated on the referred weight and height, and includes subjects with BMI >= 30; in the multivariate analysis, the reference category consists of subjects with normal weight (20=<BMI<25).

The question about smoking involved three possible answers: being a current smoker, a former smoker or having never smoked. Since we had no information on the duration of smoking or the date of quitting of former smokers, and this group can be highly heterogeneous, we decided to compare current smokers only with people who have never smoked.

Daily alcohol consumption includes beer, wine and spirits, and is treated as a dichotomous (yes/no) variable.

### Determinants and confounders

The socio-demographic characteristics analysed as possible determinants of health-related lifestyles were family composition (couples with children, couples without children, lone parents, single people and other family combinations), marital status (married living with partner, married living at distance and not married), educational level (high school or more, middle school and up to primary school) and occupational condition (occupied, job seekers and inactive).

As indicators of the migration pathway we analysed the area of birth (new EU countries, eastern Europe and Balkans, Asia, South America, North Africa and Middle East, Sub-Saharan Africa, and a mixed group of developed countries), the age at arrival in Italy (0-5, 6-13, 14-24, 25-34, 35+), the length of residence in years (0-4, 5-9, 10-14, 15+), and the reason for migration (work, family, forced , and a residual category of other reasons). We also assessed the ease of their travel to Italy, based on the type of transportation used: ‘comfortable’ whenever they report taking an airplane, ‘uncomfortable’ if the migration involved travelling on foot or by boat, and ‘average’ when they used any other means of transportation (train, car, ferry, and bus).

Finally, integration was estimated by means of two indicators: the sense of solitude in Italy and language difficulties with the doctor, obtained by combining difficulties in explaining symptoms with those in understanding therapeutic prescriptions.

As possible confounders, we included the age at the interview in 10-year age classes and four macro-regions of residence to account for possible geographical variations.

### Statistical methods

The independent impact of all the determinants on health behaviours was assessed through robust multivariable Poisson models estimating prevalence rate ratios (PRR) [10]. All the analyses were weighted by the Istat normalized survey weights and stratified by gender.

A correlation analysis verified the absence of collinearity among all variables, except for that between age, length of residence and age at arrival. To overcome this problem, length of residence and age at arrival were evaluated separately in the models including all the other variables; for each outcome, we present only the estimates derived from the model with the highest explanatory power measured by the Bayesian information criterion [11].

Results have been computed using the R and STATA statistical software [12, 13].

## Results

The distribution of determinants and outcomes in the study population is reported in Table 1. This population has a slight female dominance (56%) and consist predominantly of families of couples with children (more than 50%), although the share of single people is fairly important (21% of men and 18% of women). Married couples living at distance account for only 9% among men and 6% among women. A medium-high level of education is observed, since only 15% of men has only primary education; women have a higher percentage of high qualifications than men (45% vs. 32%) but they are less occupied (55% vs. 78%).

**Table 1.** Socio-demographic characteristics, migration pathway, indicators of integration, and health behaviours among resident immigrants aged 15-64, by gender. Italy, 2011-2012.

|  |  | Men (n=6947) |  |  | Women (n=8783) |  |  |
| --- | --- | --- | --- | --- | --- | --- | --- |
|  |  | N | %* | 95% CI | N | %* | 95% CI |
| **Socio-demographic characteristics** |  |  |  |  |  |  |  |
| Age at interview | 15-24 | 1093 | 17.2 | 16.0-18.6 | 1186 | 14.3 | 13.3-15.4 |
|  | 25-34 | 1881 | 30.2 | 28.6-31.8 | 2527 | 30.3 | 29.0-31.7 |
|  | 35-44 | 2182 | 30.6 | 29.0-32.1 | 2588 | 29.0 | 27.5-30.1 |
|  | 45-54 | 1318 | 16.9 | 15.7-18.2 | 1708 | 18.1 | 17.1-19.2 |
|  | 55-64 | 473 | 5.1 | 4.5-5.8 | 774 | 8.5 | 7.7-9.3 |
| Area of residence | North-Est | 1324 | 27.1 | 25.7-28.6 | 1600 | 26.3 | 25.0-27.5 |
|  | North-West | 1297 | 35.2 | 33.5-37.0 | 1583 | 33.9 | 32.4-35.4 |
|  | Central | 1190 | 24.8 | 23.6-26.1 | 1518 | 24.2 | 22.7-25.5 |
|  | South and Islands | 3136 | 13.6 | 12.9-14.2 | 4082 | 15.0 | 14.4-15.7 |
| Family composition | Couples with children | 3583 | 51.7 | 50.0-53.4 | 4139 | 50.0 | 48.5-51.4 |
|  | Couples without children | 871 | 9.3 | 8.4-10.2 | 1282 | 13.4 | 12.5-14.4 |
|  | Lone parents | 862 | 12.5 | 11.5-13.6 | 1523 | 16.3 | 15.4-17.5 |
|  | Single people | 1181 | 21.0 | 19.4-22.6 | 1630 | 18.4 | 17.3-19.5 |
|  | Other | 450 | 5.5 | 4.9-6.3 | 209 | 1.8 | 1.5-2.2 |
| Marital status | Married living with spouse | 3431 | 46.7 | 45.2-48.5 | 4255 | 50.7 | 49.2-52.1 |
|  | Married living at distance | 577 | 8.8 | 7.9-9.9 | 460 | 5.6 | 5.0-6.3 |
|  | Not married | 2939 | 44.3 | 42.6-46.0 | 4068 | 43.7 | 42.3-45.2 |
| Educational level | High school or more | 1995 | 32.4 | 30.8-34.0 | 3735 | 45.2 | 43.8-46.7 |
|  | Middle school | 3755 | 52.7 | 51.1-54.4 | 3958 | 43.7 | 42.3-45.1 |
|  | Up to primary school | 1197 | 14.9 | 13.7-16.1 | 1090 | 11.1 | 10.3-12.0 |
| Occupational condition | Occupied | 5433 | 78.0 | 76.5-79.3 | 4879 | 55.2 | 53.8-56.7 |
|  | Job seekers | 670 | 10.4 | 9.4-11.5 | 793 | 9.7 | 8.8-10.6 |
|  | Inactive | 844 | 11.6 | 10.6-12.7 | 3111 | 35.1 | 33.7-36.5 |
| **Migration pathway** |  |  |  |  |  |  |  |
| Area of birth | New EU countries | 1745 | 24.2 | 22.8-25.7 | 2947 | 30.0 | 28.9-31.5 |
|  | Eastern Europe and Balkans | 1738 | 22.2 | 20.9-23.5 | 2403 | 26.1 | 24.9-27.3 |
|  | North Africa and Middle East | 1338 | 18.5 | 17.2-20.0 | 1012 | 11.3 | 10.4-12.3 |
|  | Sub-Saharan Africa | 500 | 7.7 | 6.8-8.6 | 372 | 5.1 | 4.4-5.7 |
|  | Asia | 1100 | 17.2 | 15.9-18.5 | 948 | 12.4 | 11.4-13.5 |
|  | South America | 327 | 6.5 | 5.6-7.4 | 719 | 9.7 | 8.8-10.6 |
|  | Developed countries | 199 | 3.8 | 3.2-4.7 | 382 | 5.3 | 4.6-6.0 |
| Length of residence in Italy (years) | 0-4 | 1090 | 15.0 | 13.8-16.3 | 1631 | 17.8 | 16.7-18.9 |
|  | 5-9 | 2177 | 32.4 | 30.8-34.0 | 3413 | 39.0 | 37.6-40.4 |
|  | 10-14 | 1989 | 30.1 | 28.6-31.7 | 2420 | 28.8 | 27.5-30.1 |
|  | 15+ | 1691 | 22.5 | 21.2-23.9 | 1319 | 14.5 | 13.5-15.5 |
| Age at arrival in Italy (years) | 0-5 | 146 | 2.1 | 1.7-2.6 | 152 | 1.5 | 1.2-1.9 |
|  | 6-13 | 522 | 7.5 | 6.6-8.4 | 494 | 6.6 | 4.7-5.9 |
|  | 14-24 | 2557 | 41.3 | 39.6-42.9 | 2926 | 36.7 | 35.3-38.1 |
|  | 25-34 | 2453 | 33.7 | 32.1-35.3 | 2805 | 31.1 | 29.8-32.4 |
|  | 35+ | 1269 | 15.5 | 14.4-16.7 | 4179 | 25.4 | 24.2-26.7 |
| Reason for migration | Work | 4660 | 65.1 | 63.4-66.6 | 4179 | 45.2 | 43.8-46.6 |
|  | Family | 1662 | 25.5 | 24.1-27.0 | 3981 | 47.5 | 46.1-48.9 |
|  | Forced | 353 | 4.6 | 4.0-5.4 | 184 | 1.9 | 1.6-2.3 |
|  | Other | 270 | 4.8 | 4.0-5.6 | 439 | 5.4 | 4.8-6.1 |
| Type of transportation | Comfortable | 2781 | 46.7 | 45.0-48.4 | 3801 | 49.7 | 48.2-51.1 |
|  | Average | 3308 | 44.5 | 42.8-46.1 | 4668 | 48.1 | 46.6-49.5 |
|  | Uncomfortable | 776 | 8.8 | 8.0-9.7 | 279 | 2.3 | 1.9-2.8 |
| **Indicators of integration** |  |  |  |  |  |  |  |
| Sense of solitude in Italy | High | 1038 | 15.1 | 13.9-16.4 | 1436 | 16.2 | 15.2-17.3 |
|  | Low | 1909 | 24.5 | 23.1-26.0 | 2698 | 28.3 | 27.0-29.6 |
|  | Not any | 4000 | 60.4 | 58.7-62.0 | 4649 | 55.5 | 54.0-56.9 |
| Language difficulties with the doctor | Many | 746 | 10.9 | 9.9-12.0 | 1063 | 13.4 | 12.4-14.5 |
|  | Some | 1911 | 26.4 | 24.9-27.9 | 2316 | 24.1 | 22.9-25.4 |
|  | Not any | 3857 | 62.7 | 61.1-64.4 | 5077 | 62.5 | 61.1-63.9 |
| **Health behaviours** |  |  |  |  |  |  |  |
| Body Mass Index (only 18+ years) | Underweight (BMI<20) | 47 | 0.8 | 0.5-1.1 | 418 | 5.7 | 5.0-6.5 |
|  | Normal weight (20<BMI<25) | 3434 | 51.4 | 49.7-53.1 | 5515 | 63.5 | 62.1-64.9 |
|  | Overweight (25<BMI<30) | 2683 | 40.3 | 38.6-42.0 | 1995 | 23.2 | 22.0-24.4 |
|  | Obese (BMI>30) | 483 | 7.5 | 6.7-8.4 | 607 | 7.5 | 6.8-8.4 |
| Daily alcohol consumption | Yes | 1899 | 28.0 | 26.5-29.5 | 759 | 9.4 | 8.6-10.3 |
|  | No | 5048 | 72.0 | 70.5-73.5 | 8024 | 90.6 | 89.7-91.4 |
| Smoking habit | Current smoker | 2390 | 33.6 | 32.0-35.2 | 1553 | 15.8 | 14.8-16.9 |
|  | Former smoker | 1110 | 15.4 | 14.3-16.6 | 981 | 11.0 | 10.1-12.0 |
|  | Never smoker | 3447 | 51.0 | 49.3-52.7 | 6249 | 73.2 | 72.9-74.4 |

\* percentages do not correspond to the Ns because they are weighted by a weight that accounts for the sampling design

Looking at the migration pathway, we observed that more than half of the population arrives from Eastern Europe and from the new EU countries, mostly women (56% vs. 46%), while about 20% arrives from Africa, but in this case the gender distribution is inverse (26% among men vs. 16% among women). There is also a small proportion of immigrants from developed countries (about 4%), which were not considered in the following multivariate analyses. Only 17% has lived in Italy for less than 5 years. Women arrived later (14% has a length of residence longer than 14 years vs. 22% among men) and at an older age (56% was older than 24 years on arrival vs. 49% among men). The reasons for migration are also very different between men and women: 65% of men and 45% of women have moved to look for a job, while 26% of men and 48% of women for family reasons. Forced migration is not very common within this resident population (5% and 2%, respectively). The majority of respondents have travelled by plane for at least a part of their journey, while an uncomfortable trip is almost 4 times more frequent among men than among women.

More than 55% of respondents state that they do not feel alone nor they have had any problem in communicating with doctors.

All unhealthy life styles are more common among men than among women: in fact, almost half of the male population is overweight or obese compared to one third of women; 28% make daily alcohol consumption (9% among women) and one third is a regular smoker (16% among women).

Multivariable results are presented in Figs 1-3, where for each outcome we show only the variables with at least one statistically significant PRR. The results of the complete models are presented in the supplementary S1-S3 Tables.

**Fig 1.**
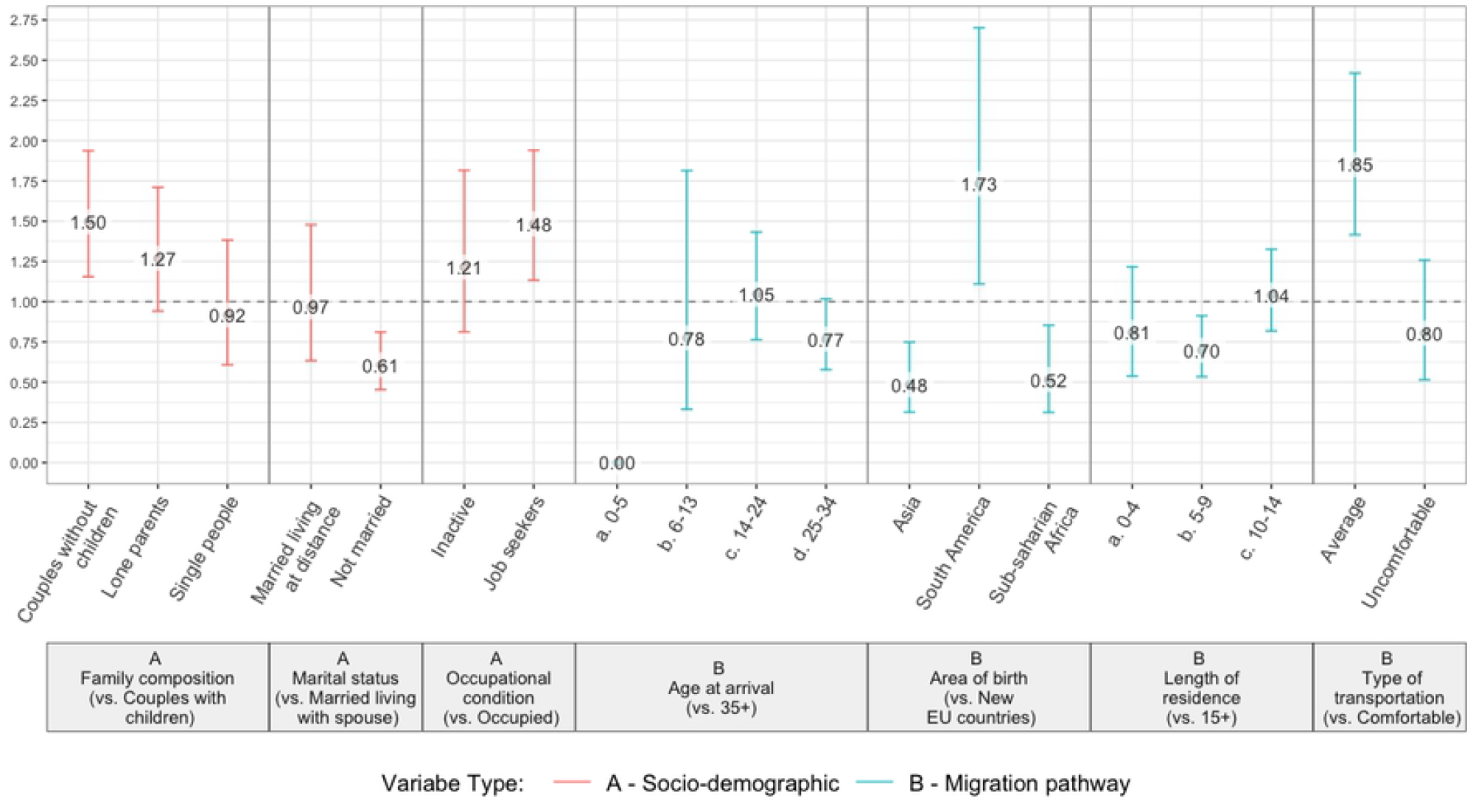
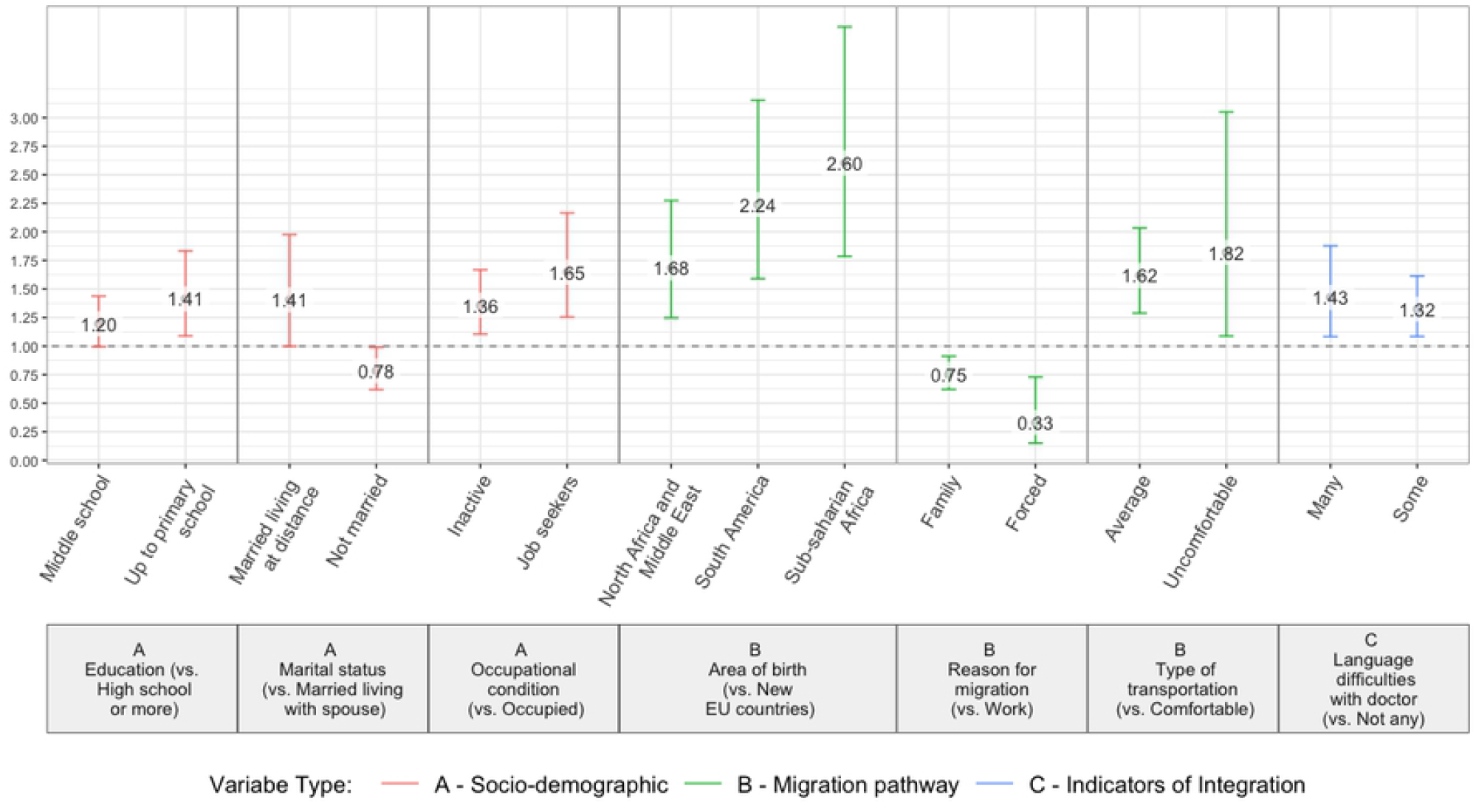
Obesity (BMI>30). Prevalence Rate Ratios (with 90% CI) of statistically significant determinants (*). Men (a) and women (b) (*) estimates from robust Poisson models, mutually adjusted for all the variables in Table 1 (length of residence and age at arrival evaluated separately)

**Fig 2.**
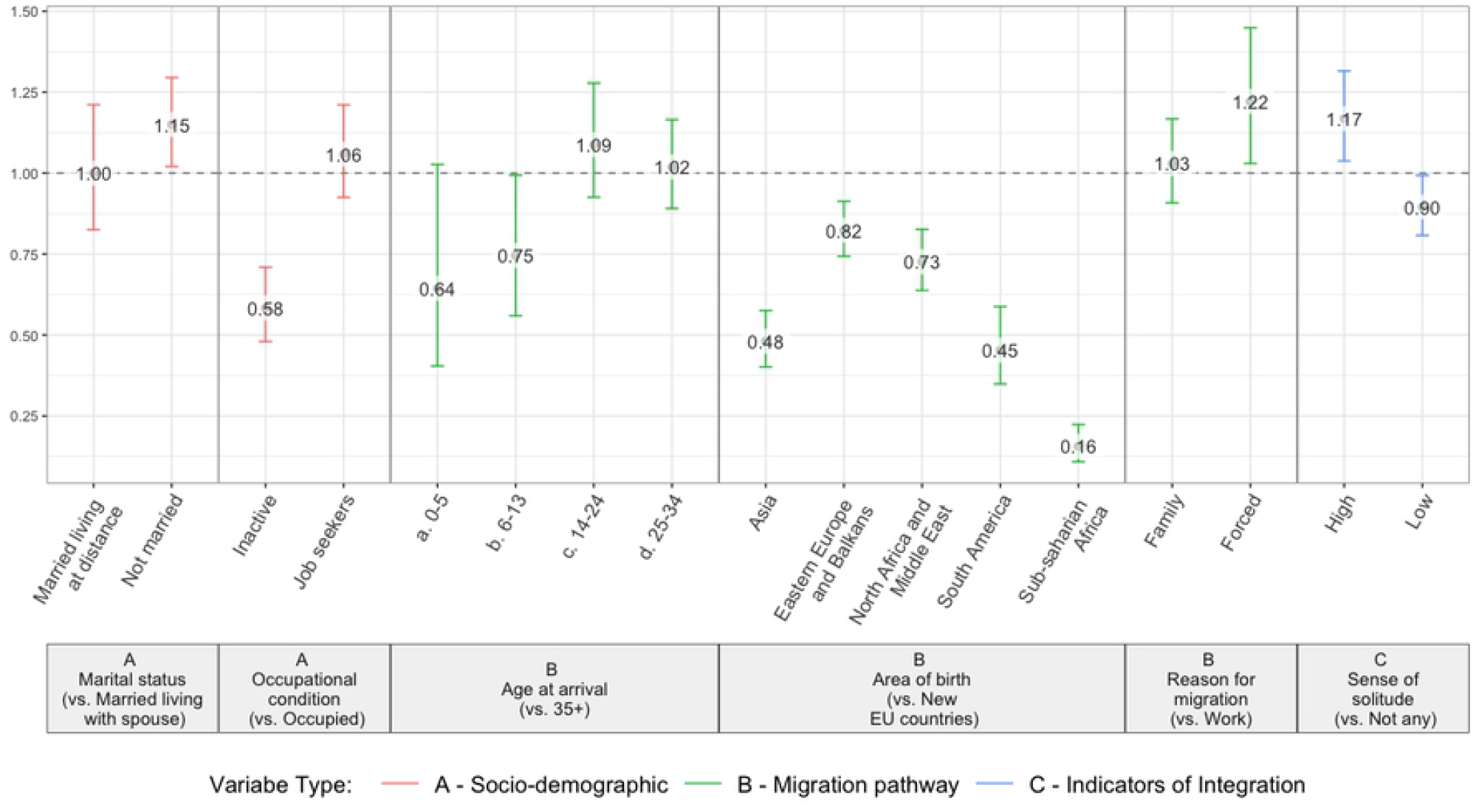
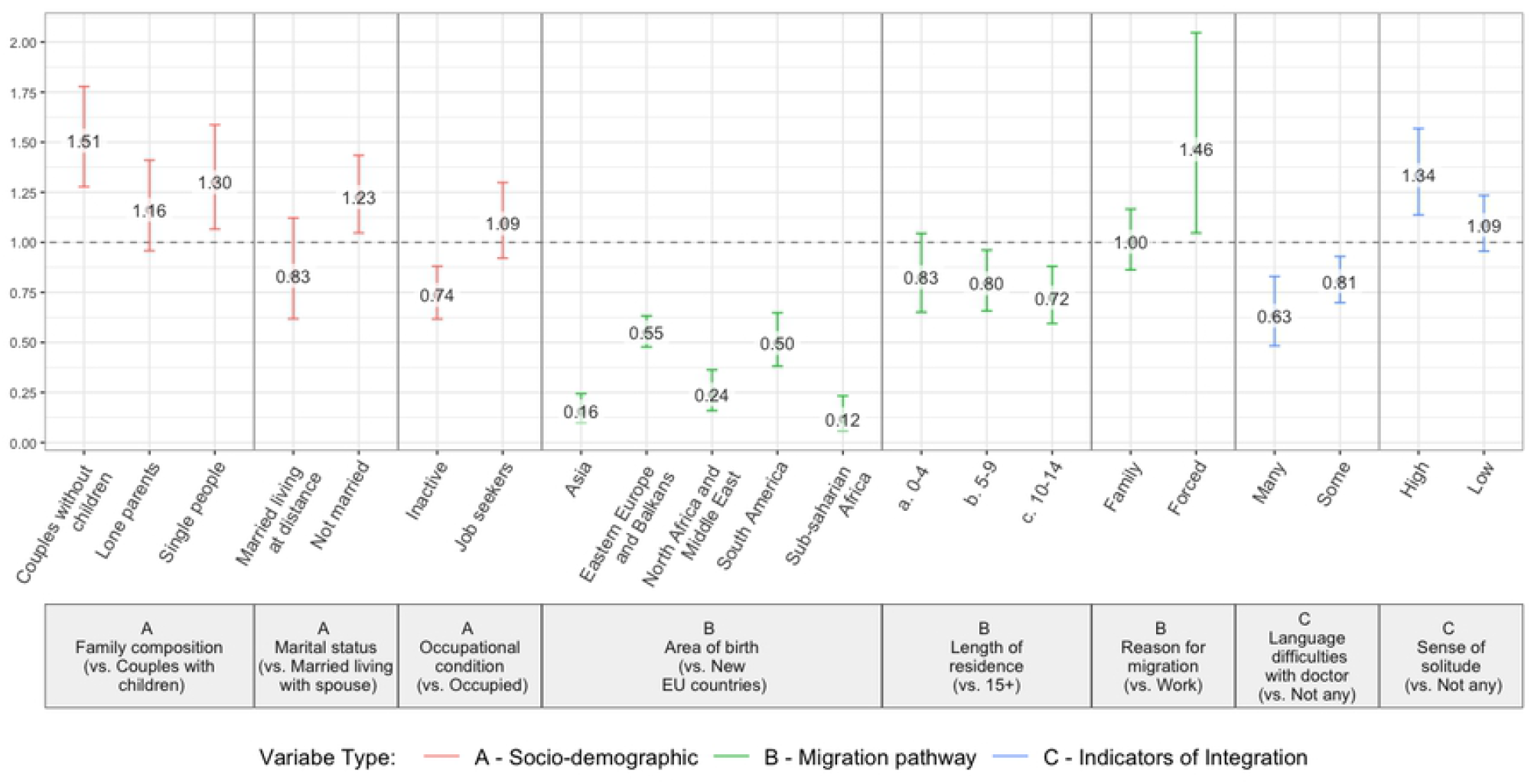
Current smoking. Prevalence Rate Ratios (with 90% CI) of statistically significant determinants (*). Men (a) and women (b) (*) estimates from robust Poisson models, mutually adjusted for all the variables in Table 1 (length of residence and age at arrival evaluated separately)

**Fig 3.**
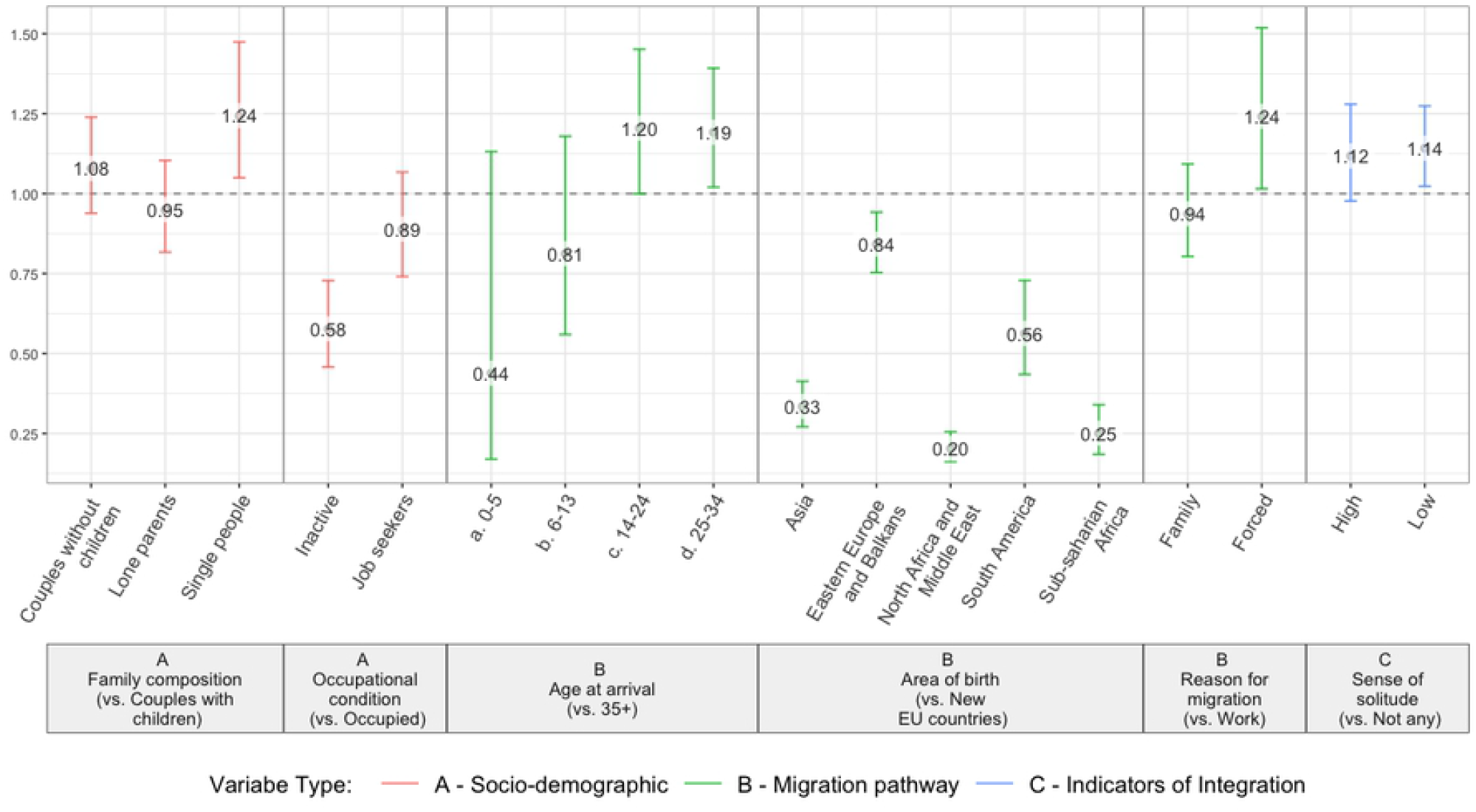
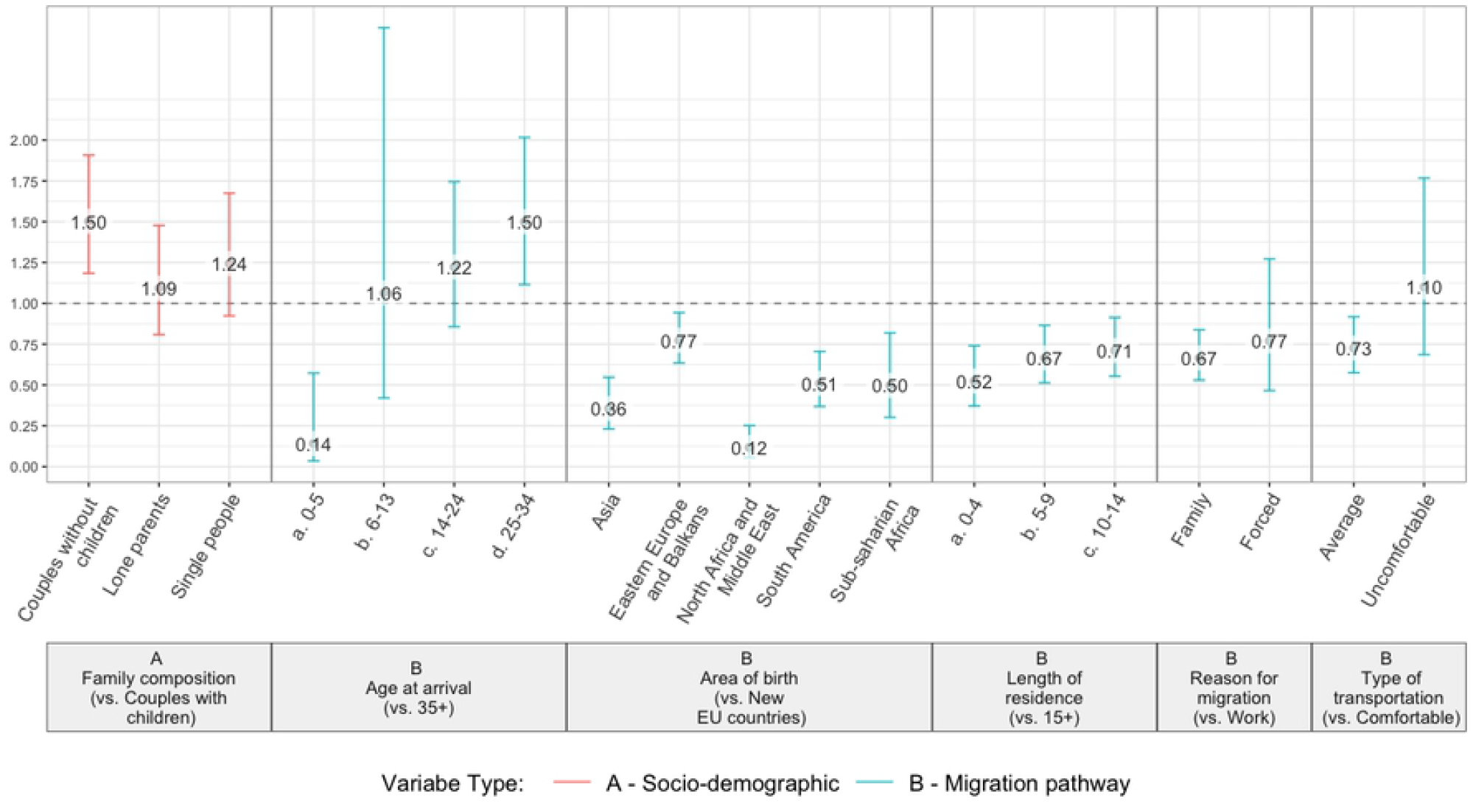
Daily consumption of alcohol. Prevalence Rate Ratios (with 90% CI) of statistically significant determinants (*). Men (a) and women (b) (*) estimates from robust Poisson models, mutually adjusted for all the variables in Table 1 (length of residence and age at arrival evaluated separately)

As regards the socio-demographic characteristics, couples without children compared to those with children are at higher risk of obesity among men (Fig 1a) and of smoking and drinking alcohol among women (Figs 2b and 3b), with excess risks generally exceeding 50%. Not married people or those living alone show lower but still significant excesses of risk; only not married men appear protected from obesity (PRR=0.61, Fig 1a). People seeking job have a greater prevalence of obesity (Fig 1), while inactive ones are protected from smoking and alcohol (Figs 2 and 3). A low educational level, instead, is significantly associated only with an increased risk of obesity among women (PRR=1.41, Fig1b).

Looking at the migration pathway, non-European citizens always show healthier behaviours, except for African women and South American men and women, who are more likely to be obese than Europeans (Fig 1). A longer duration of residence in Italy is associated with an increased probability of acquiring unhealthy behaviours among women. Furthermore, alcohol consumption appears to be associated also with younger ages at arrival, with an excess of risk of about 50% among women aged 25-34 (Fig 3b) and 20% among men aged 14-34 (Fig 3a). The reason for migration also plays an important role in explaining lifestyles, particularly among women: those arriving for family reasons are 25% less likely to be obese (PRR=0.75, Fig 1b) and 33% less likely to drink alcohol (PRR=0.67, Fig 3b) than those who arrive for work. Women declaring a forced migration are also protected from obesity, but at greater risk of smoking (PRR=1.46, Fig 2b), similarly to men (PRR=1.22, Fig 2a). Moreover, men declaring a forced migration are more exposed to alcohol consumption (PRR=1.24, Fig 3a). On the other hand, compared to people travelling by plane, men and women with a less comfortable trip have a higher probability of obesity (Fig 1).

Finally, among resident immigrants we observe also a significant impact of the integration indicators: the sense of loneliness increases the risk of smoking by 34% among women and 17% among men (Fig 2) and the risk of daily alcohol by 14% among men (Fig 3a). Women who report many difficulties in communicating with their doctor are more likely to be obese (PRR=1.43, Fig 1b), but appear protected from smoking (PRR=0.63, Fig 2b).

## Discussion

The survey involved more than 15000 first generation immigrants, most of whom arrived from European countries and with a medium-long duration of residence in Italy. As expected, forced migration is not very common within this resident population, who on average declares quite good levels of integration, as measured by the sense of loneliness and language difficulties. Nonetheless, the profile of unhealthy lifestyles highlights important variations within this population, which should be acknowledged and taken into account for the development of effective preventive strategies.

Unhealthy lifestyles are more common among men than among women and vary widely by ethnic group, with most non-European migrants less at risk than Europeans. Cultural factors may account for some of the observed variations. The observance of the Muslim religious prohibition of drinking alcoholics may substantially explain the low alcohol consumption rates observed in our study, as well as in other European countries [14, 15, 16, 17], and the significant advantage of immigrants arriving from non-European countries.

The high prevalence of obesity among South American immigrants could be related to the nutritional transition that occurred in the early 2000’s in Latin-American countries, where immigrants may have been exposed to an unhealthy diet and have increased their risk of obesity even before leaving their country [18]. The social dimension of food, with high expectations for a rich and fat Western diet as opposed to the healthier traditional diet [19], may instead be responsible for the excess risk of obesity of African women. Furthermore, qualitative studies emphasize the fatalistic approach to health of some ethnic minorities, particularly from sub-Saharan Africa, which implies low awareness of unhealthy behaviours [20]. Cultural and socioeconomic factors are also often responsible for low levels of physical activity, which in turn reflect on obesity, and which is often perceived as a luxury rather than a need for health, particularly among females [21, 22, 23].

Looking at the socio-demographic characteristics analysed in our study, a common trait is represented by the significant impact of employment status and family composition, while the educational level loses significance. People seeking job and single people are generally at higher risk of unhealthy lifestyles. Immigrants in fact tend to share characteristics of vulnerability and socioeconomic deprivation with the lowest classes of the host population, which are well known determinants of poor health and unhealthy behaviours [24, 25]. Low socioeconomic conditions represent therefore an additional threat for immigrants’ health that cannot be overlooked by health prevention strategies [6, 26, 27].

A longer duration of residence in Italy is associated with an increase in risky behaviours. This is in line with the literature suggesting that the process of acculturation of migrants in the host country often brings the acquisition of unhealthy lifestyles, generally less common in the countries of origin [28, 6]. In our data this phenomenon is particularly evident among women. The acculturation process naturally evolves with time from arrival and is usually faster among people more in contact with the host population [7], consistently with our results showing higher risks among immigrants arrived in a working age or for work reasons. Conversely, inactive people and women that have arrived for family reasons, who are more likely to be outside the labour market and less in contact with the host population, and therefore subject to a slower acculturation process, resulted protected from alcohol and smoking habits.

Not surprisingly, a forced migration is at higher risk of smoking and alcohol consumption, as a response to the stress originating from this life experiences [29]. A similar mechanism could explain the excess of obesity among women who travelled to Italy less comfortably than by plane.

As mentioned before, immigrants who have been interviewed have not arrived in Italy very recently. Consequently, very few respondents state that they feel alone or have problems in communicating with doctors. However, their proportion is still relevant, particularly among women, who are more frequently inactive and therefore less likely to have a satisfying network of social relationships. Among them, in fact, we observed also a significant impact of these integration indicators, which seem less important for men.

The main limitation of this study is that the survey included only the resident population, who had a medium-long duration of stay in Italy. Therefore, respondents represent the more integrated part of the foreign population and the results cannot be generalised to newly arrived undocumented migrants. Resident foreigners are however the majority of the immigrant population in Italy, equalling 8-9% of the total resident population, while the percentage of undocumented migrants is estimated to be less than 1% [30]. Furthermore, we have studied the independent effect on unhealthy lifestyles of a great variety of factors, but we do not yet know how they interact and modulate the behavioural risks. More research is needed in order to identify the combination of factors that constitute the subgroups of the immigrant population most in need of preventive strategies.

## Conclusions

The profile of the main unhealthy lifestyles among migrants is shaped by cultural, socioeconomic and migratory characteristics. Important differences have been observed between men and women and between countries of origin. Understanding these factors can help to design more tailored preventive messages, which are needed to interrupt the deterioration of the migrants’ health capital. Low levels of integration can have an additional negative impact on health and more inclusion and integration policies should complement health promotion strategies. Future research will specifically address which aspect of the lack of integration in the host country might be most associated with the uptake of unhealthy behaviours.

## Data Availability

We used unidentifiable data derived from a survey conducted by the National Institute of Statistics (Istat), which keeps the property of the data. Their use has been authorized under a specific license for the project Immigration, integration, settlement. Italian-Style, funded by the Italian Ministry for Education, University and Research, therefore we are not authorized to share the data with third parties. Data are however available from Istat itself upon reasonable request, by making application to this website: https://contact.istat.it/index.php?Lingua=Inglese. Istat have carried out the necessary procedures for the approval of the ethics committee and provided anonymous data, which have been processed following appropriate privacy regulations study characteristics and results are reported according to the STROBE statement.

https://www.istat.it/it/archivio/191090

## Acknowledgements

The authors wish to thank Prof. Viviana Egidi and Dr. Silvia Loi for fruitful discussions on the methodology during the first stages of this work. The analyses here presented are part of the project “ Immigration, integration, settlement. Italian-Style”, financed by the Italian Ministry for Education, University and Research.

